# Cumulative TB disease burden following sputum Xpert Ultra ‘Trace’ results in clinical settings: Results from a multi-site observational clinical study

**DOI:** 10.1101/2025.07.14.25331384

**Authors:** Ronit R Dalmat, Caitlin Visek, Elvira Budiawan, Gabrielle Stein, Annet Nalutaaya, James Mukiibi, Mariam Nantale, Patrick Biché, Joowhan Sung, Zanele Magcaba, Nompumelelo Ngcobo, Jennifer F. Morton, Meena Lenn, Samantha Aucock, Adrienne E. Shapiro, Amy Steadman, Robin Draper, Amanda Ganguloo, Douglas Wilson, Achilles Katamba, Emily A Kendall, Paul K Drain

## Abstract

**Background:** Highly sensitive molecular tests, like Xpert Ultra, are reshaping TB diagnosis—detecting paucibacillary TB but sometimes creating uncertainty when they detect DNA in extremely low quantities that may not signal disease. This ambiguity also complicates the evaluation of novel diagnostic strategies. We sought to monitor adults with a ‘Trace’ result on an Xpert Ultra test to estimate the risk of tuberculosis disease up to 24 months later.

**Methods:** We conducted a multi-site clinical observational study in South Africa and Uganda, where we enrolled ambulatory participants aged >=15 years with a sputum Xpert Ultra Trace result who had not yet initiated TB treatment. All participants underwent comprehensive clinical evaluation and repeated, standard sputum TB testing. Clinicians deferred treatment recommendations if TB status remained uncertain after evaluation. Untreated participants were followed regularly until TB diagnosis and/or treatment initiation. We estimated cumulative incidence of TB disease, defined by three reference standards.

**Results:** Of 311 participants with Trace results (50% male, 57% PLHIV, 37% treated for TB within the last 5 years), 24% were positive for TB within 12 months by culture, 37% by sputum Xpert or culture, and 54% by culture or clinical diagnosis. After excluding those diagnosed with TB at baseline, patients identified at baseline as having recent TB history, abnormal chest x-ray, or positive tongue swab, had higher risk of TB diagnosis (hazard ratios: 2.6, 2.4, 4.5, respectively) during follow-up. This hazard was highest in the first three months after the negative baseline evaluation (0.22 [95% CI: 0.19-0.26] per person-month) and decreased to 0.01 [95% CI: 0-0.02] per person-month in both the 3-6 and 6-12-month intervals.

**Conclusion:** Approximately half of adults and adolescents with a sputum Trace result were diagnosed with TB disease within twelve months. Although most TB diagnoses were made within 3 months, risk remained higher than estimated population incidence rates through the follow-up period. Individuals with sputum Trace results should receive close clinical monitoring, despite initial clinical treatment decision.

**Summary results:** Xpert Ultra “Trace” results detect low quantities of TB DNA, creating diagnostic uncertainty that complicates both clinical decisions and diagnostics evaluation. This clinical observational cohort study followed untreated patients with Trace results for twelve or more months, using survival analysis to estimate actual tuberculosis disease risk and identify risk factors.

## Introduction

Nearly 11 million people developed tuberculosis (TB) disease in 2023,^1^ and TB remains the leading infectious cause of death. Delayed diagnosis is a major contributor to TB morbidity and mortality. Molecular TB diagnostic tests substantially improve sensitivity over sputum smear microscopy^2^, and their wider adoption could improve detection of TB at early and paucibacillary stages, leading to earlier treatment that may reduce transmission and prevent severe disease outcomes. However, the small quantities of *Mycobacterium tuberculosis* (Mtb) DNA detected by molecular tests can sometimes be residual from prior TB,^3–6^ and for individuals who lack other microbiological evidence of current disease, the significance of small quantities of Mtb DNA remains unclear^4^. One such result is the diagnostic readout of “Trace MTB detected” from GeneXpert’s current TB diagnostic cartridge, Xpert MTB/RIF Ultra (“Ultra”).^6^ This readout indicates presence of Mtb-specific *IS6110* and/or *IS1081* molecular signals (cycle threshold [Ct]<37), but without a signal from *rpoB* probes^7^.

WHO guidelines^8^ recommend that treatment decisions for patients with Xpert Ultra Trace results be based on clinical presentation and patient context (including prior TB history and probability of relapse), and do not recommend routine repeat testing in adults with pulmonary TB symptoms. Among people with sputum Trace results in recent studies, the proportion with culture-confirmed TB disease varied considerably, ranging from 8-52%^9–15^.

We sought to better understand the burden of disease associated with a Trace result on a sputum Ultra test, including disease that is not readily confirmable with reflexive sputum testing (e.g., culture or repeat Ultra), on routine testing in high-burden clinical settings. We previously reported that 20% of patients with a Trace Ultra result had positive sputum cultures and a total of 39% were diagnosed with TB disease upon initial evaluation,^16^ with an additional 8% diagnosed with TB disease (including 2% who were newly culture positive) within three months. We now extend the follow-up period to assess the ongoing risk of TB disease beyond those early clinical evaluations, aiming to estimate the 6- and 12-month cumulative incidence and identify risk factors for a TB diagnosis, both cumulatively and among those not diagnosed at baseline.

## Methods

### Study design, participants, and procedures

We conducted a prospective, observational cohort study of patients who received a Trace result on an expectorated sputum Xpert Ultra test conducted as part of routine clinical care. Patients were recruited from seven health facilities in Kampala, Uganda, between February 2021 and July 2024, and from 35 outpatient clinics in the catchment area of Harry Gwala Regional Hospital in Pietermaritzburg, South Africa, between November 2022 and June 2024. Patients were eligible for recruitment if they were aged >=15 years in Uganda or >=16 years in South Africa, had not yet initiated TB treatment at the time of the enrollment (“baseline”) visit, did not require hospitalization, and had not received any TB treatment within the previous three months.

Within 14 days of their Trace positive result, consenting participants underwent an extensive baseline evaluation, which comprised a standardized interview (including history of prior TB diagnosis and treatment, current symptoms, and other comorbidities), physical exam with vital signs, chest x-ray (CXR) with radiologist interpretation, repeat sputum Ultra testing, sputum cultures (liquid and solid culture on each of two expectorated specimens), HIV testing (followed by CD4 count, as indicated), and serum C-reactive protein (CRP: *LumiraDx*, South Africa; *ichroma*, Uganda) measurement. All participants in South Africa and participants with HIV in Uganda also underwent urine lipoarabinomannan (*Abbott Determine LAM*) testing. All participants in Uganda underwent a chest computed tomography scan at baseline. Additionally, two tongue swabs (*Copan FLOQSwab*) were collected per a previously published protocol^17^ and analyzed on an investigational qPCR assay^18^. With the exception of tongue swabs (which were analyzed retrospectively), results of this baseline evaluation were made available to clinicians for treatment-related decision-making.

Participants who remained untreated following baseline evaluation were reassessed until they received a diagnosis of TB and initiated treatment, with the following visit schedule: one, three, six, nine, twelve, and (*in Uganda only*) twenty-four months (Table of procedures: **Table S-1**).

### Monitoring and treatment initiation

Participants enrolled in Uganda remained under the care of TB clinicians at the referring health facilities, who could start treatment at any time but were encouraged to await study investigations when TB status was uncertain. An advisory panel of pulmonologists and radiologists additionally reviewed new study data and made treatment recommendations accordingly.

For participants enrolled in South Africa, a TB physician evaluated each participant at each visit and could decide to make a diagnosis of TB and initiate treatment. A committee of three TB physicians determined whether to recommend deferral of TB treatment.

Participants who initiated treatment were treated according to the standard of care and followed on a different research visit schedule for outcomes.

### Outcome definitions

The outcome of interest was TB disease as defined by each of three reference standards: composite, microbiological, and culture only. By the composite reference standard (CRS), TB was defined as having any Mtb-positive sputum culture result (either liquid or solid), a recommendation to start TB treatment, or a death attributed to TB. The microbiological reference standard was defined as having either an Mtb-positive sputum culture result or a positive sputum Ultra with a semiquantitative result of ‘Very Low’ or higher. The culture-only standard defined TB as having any Mtb-positive sputum culture result.

### Missing data

In cases where the date of treatment recommendation was not available, whether due to missing records or treatment that was started outside of a study visit, the date of treatment initiation was used in its place. Anyone with a missing covariate value was excluded from analyses involving that covariate because no single variable had >10% missingness (*a priori* threshold).

### Statistical analysis

Cumulative incidence of TB diagnosis (using three TB reference standards) was estimated at 6, 12, and 24 months for the full cohort, beginning at enrollment. In a secondary analysis, cumulative incidence of TB diagnosis was estimated among only those individuals who had a negative TB status according to the CRS at baseline (termed the “follow-up cohort”).

Cumulative incidence of TB diagnosis, as defined by the CRS, was estimated using Kaplan-Meier (KM) estimation, with follow-up censored at times of non-TB deaths or at the end of available follow-up (last attended visit date due to conclusion of visit schedule, loss to follow-up, or data availability at time of analysis). For MRS- and culture-defined TB, cumulative incidence was estimated using the Aalen-Johansen estimator to account for competing events (initiation of TB treatment or TB-related death prior to microbiological or culture confirmation); as for the CRS definition, follow-up time was censored when participants died of non-TB causes or reached the end of available follow-up. Participants who were recommended treatment at baseline, or during a follow-up visit, without MRS or culture confirmation but declined to initiate therapy were counted as remaining at risk for MRS- and culture-defined TB outcomes. Hazard rates were also calculated within consecutive time periods to compare the instantaneous TB risk per unit time at different intervals since a Trace result.

We compared cumulative cause-specific hazards of MRS-positive TB by a set of baseline characteristics and test results using Cox proportional hazards regression models: HIV status (stratified by CD4 count < or >200 cells/mm^3^), history of prior TB (< 5 years, >5 years, none), site (South Africa vs. Uganda), age (<35 vs. >35 years), sex (male vs. female), CXR result (normal vs. abnormal, as determined by radiologists), and tongue swab result (either of two swabs positive vs. both negative). Time was measured from baseline until microbiological confirmation of TB or censoring (death, treatment, or end of follow-up). Hazard ratios were calculated for each characteristic comparison, along with corresponding 95% confidence intervals.

### Ethical approvals

The study was approved by the Makerere University School of Public Health Research and Ethics Committee, the University of KwaZulu-Natal Biomedical Research Ethics Committee, and the institutional review boards at the University of Washington and Johns Hopkins University.

## Results

### Cohort characteristics: overall and follow-up cohort

A total of 311 adults and adolescents (170 in Uganda and 141 in South Africa) were enrolled with a Trace result on routine clinical TB testing on Ultra. Of these participants, 191 (61%) had no evidence of TB (per CRS) at baseline and were included in the follow-up cohort (**Figure 1**). This follow-up cohort had median age 38 years [IQR: 30-48], 43% were male, and 44% reported prior TB treatment (**Table 1**). Half (52%) were people living with HIV, nearly all of whom (86%) had CD4 counts >200 cell/mm^3^ at baseline. At baseline, most of this follow-up cohort had at least one TB-related symptom (82%), and fewer than half (40%) had ‘normal’ chest x-ray findings. At time of analysis, all participants had the opportunity for at least six months of follow-up; a subset (N=35) had completed 24 months of follow-up.

**Table 1.** Baseline demographics of Trace cohort and subset of CRS-negative in the follow-up cohort.

| **Characteristics** |  | **Trace cohort** |  | **Trace follow-up cohort** |  |  |  |  |  |
| --- | --- | --- | --- | --- | --- | --- | --- | --- | --- |
|  |  | Total Enrollment<br>(n=311) |  | CRS- at Baseline<br>(n=191) |  | TB positive *beyond* baseline |  |  |  |
|  |  |  |  |  |  | MRS+<br>(n=28) |  | CRS+<br>(n=34) |  |
|  |  | n | % | n | % | n | % | n | % |
| **Country** | Uganda | 170 | (54.7%) | 91 | (47.6%) | 9 | (32.1%) | 8 | (23.5%) |
|  | South Africa | 141 | (45.3%) | 100 | (52.4%) | 19 | (67.9%) | 26 | (76.5%) |
| **Age, years (median [IQR])** |  | 37 | [30-47] | 38 | [30-48] | 39 | [32-51.8] | 39.5 | [32.8-55.8] |
| **Sex** | Male | 154 | (49.5%) | 83 | (43.5%) | 19 | (67.9%) | 17 | (50.0%) |
| **Symptoms** | Cough | 243 | (78.1%) | 140 | (73.3%) | 21 | (75.0%) | 25 | (73.5%) |
|  | Fever | 118 | (37.9%) | 61 | (31.9%) | 9 | (32.1%) | 10 | (29.4%) |
|  | Weight Loss | 146 | (46.9%) | 70 | (36.6%) | 13 | (46.4%) | 15 | (44.1%) |
|  | Night Sweats | 88 | (28.3%) | 42 | (22.0%) | 6 | (21.4%) | 11 | (32.4%) |
|  | None | 40 | (12.9%) | 35 | (18.3%) | 4 | (14.3%) | 4 | (11.8%) |
| **Underweight** |  | 52 | (16.7%) | 25 | (13.1%) | 3 | (10.7%) | 4 | (11.8%) |
| *Missing* |  | 3 | (1.0%) | 0 | (0%) | 0 | (0%) | 0 | (0%) |
| **HIV status** | Positive | 178 | (57.2%) | 99 | (51.8%) | 14 | (50.0%) | 26 | (76.5%) |
|  | CD4<=200/mm<sup>3</sup> | 35/178 | (19.7%) | 12/99 | (12.1%) | 4/14 | (28.6%) | 8/26 | (30.8%) |
|  | *Missing* | 10/178 | (5.6%) | 2/99 | (2.0%) | 0/1 | (0%) | 0/26 | (0%) |
| **Alcohol use** | Positive AUDIT-C score<sup>1</sup> | 106 | (34.1%) | 58 | (30.4%) | 11 | (39.3%) | 11 | (32.4%) |
| **Pregnant** |  | 11/157 | (7.0%) | 8/108 | (7.4%) | 2/9 | (22.2%) | 2/17 | (11.8%) |
| *Missing* |  | 1/157 | (0.6%) | 0/108 | (0%) | 0/9 | (0%) | 0/17 | (0%) |
| **Prior TB treatment (ever)** |  | 114 | (36.7%) | 84 | (44.0%) | 17 | (60.7%) | 14 | (41.2%) |
| **Treated within the last 5 years** |  | 72/114 | (63.2%) | 58 | (69.0%) | 14 | (82.4%) | 10 | (71.4%) |
| *Missing* |  | 1 | (0.3%) | 0 | (0%) | 0 | (0%) | 1 | (7.1%) |
| **Recent household contact with TB<sup>2</sup>** |  | 25 | (8.0%) | 17 | (8.9%) | 2 | (7.1%) | 1 | (2.9%) |
| **Elevated CRP (>= 5 mg/L)** |  | 155 | (49.8%) | 75 | (39.3%) | 19 | (67.9%) | 21 | (61.8%) |
| *Missing* |  | 1 | (0.3%) | 0 | (0%) | 0 | (0%) | 0 | (0%) |
| **Chest x-ray result** | Normal | 106 | (34.1%) | 77 | (40.3%) | 6 | (21.4%) | 5 | (14.7%) |
|  | Cavitary lesion | 24 | (7.7%) | 14 | (7.3%) | 3 | (10.7%) | 0 | (0%) |
|  | Other abnormality | 172 | (55.3%) | 93 | (48.7%) | 19 | (67.9%) | 28 | (82.4%) |
|  | *Missing* | 9 | (2.9%) | 7 | (3.7%) | 0 | (0%) | 1 | (2.9%) |
| **Repeat sputum Xpert Ultra result** | High | 0 | (0%) | 0 | (0%) | 0 | (0%) | 0 | (0%) |
|  | Medium | 2 | (0.6%) | 0 | (0%) | 0 | (0%) | 0 | (0%) |
|  | Low | 25 | (8.0%) | 5 | (2.6%) | 1 | (3.6%) | 1 | (2.9%) |
|  | Very low | 25 | (8.0%) | 5 | (2.6%) | 0 | (0%) | 2 | (5.9%) |
|  | Trace | 35 | (11.3%) | 14 | (7.3%) | 6 | (21.4%) | 6 | (17.6%) |
|  | Negative | 215 | (69.1%) | 159 | (83.2%) | 18 | (64.3%) | 23 | (67.6%) |
|  | *Missing* | 9 | (2.9%) | 8 | (4.2%) | 3 | (10.7%) | 2 | (5.9%) |

<sup>1</sup>Score of 3 or greater for women, or 4 or greater for men. <sup>2</sup>Participant was living, at baseline, with someone who was diagnosed with active TB in the past 6 months. \*Follow-up ongoing for some participants, expected completion early 2026; % calculated based on column total (N) unless otherwise specified n/N

**Figure 1.**
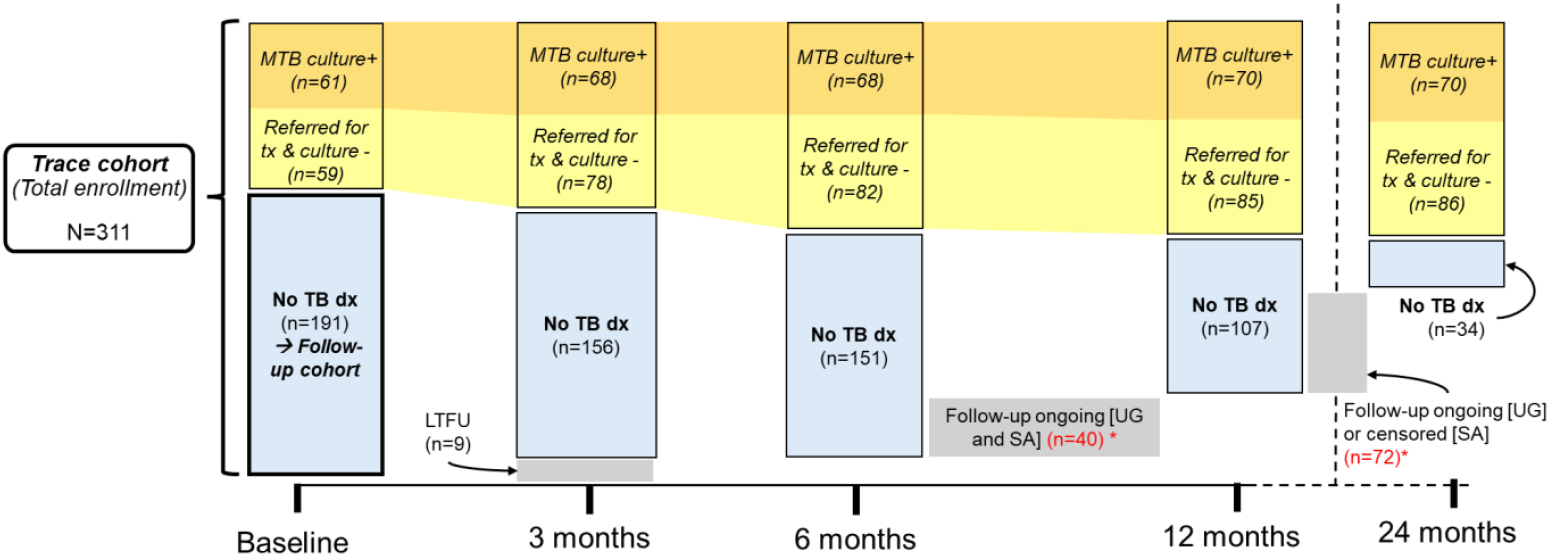
Flow chart of Trace (total enrollment) and follow-up cohorts analyzed. Classifications were made with respect to test results and follow-up visits that occurred through March 31, 2025. Number of participants with a positive Mtb culture (solid or liquid) at each time point are orange; yellow is the number who were culture-negative but referred for treatment. Thus the sum of orange and yellow boxes represents the cumulative incidence of TB by CRS at each time point. The Trace cohort includes all participants enrolled (N=311) followed for outcomes; the follow-up cohort comprises the subset of participants who did not have TB (per CRS) after baseline evaluation and testing. Gray boxes show the number of participants whose follow-up was censored due to incomplete follow-up (the cohort is undergoing follow-up for at least 12 months after baseline so visits are projected through March 2026). Participants enrolled in Uganda (UG) by August 2024 are undergoing an additional visit at 24 months; follow-up for all participants enrolled in South Africa (SA) concluded at 12 months.

### Cumulative incidence of diagnoses from baseline: Total enrollment

Among all participants with a Trace result, cumulative incidence of TB by the CRS definition was 49.0% (95% CI: 43.4-54.7%; n=150 events) at 6 months, and 53.5% (95% CI: 46.9-60.1%; n=155) at 12 months. Determined by the much smaller subset of individuals from Uganda who were followed beyond 12 months, incidence was slightly higher at 24 months: 54.9% (95% CI: 47.9-61.9%; n=156) (**Table 2**). Incidence of microbiologically confirmed TB was 32.6% (95% CI: 27.4–37.8%; n=100) and 37.3% (95% CI: 31.1% - 43.5%; n=106) at 12 months. There were no microbiologically confirmed TB diagnoses between 12-24 months of follow-up. Incidence of TB by the culture definition was less than half that estimated by the CRS: 22.0% (95% CI: 17.4-26.6%; n=68) at 6 months and 23.6%, 95% CI: (95% CI: 18.5-28.7%; n=70) at 12 months. Across all reference standards, diagnoses were made at the highest rate in the first 30 days (**Figure 2a-c**).

**Table 2.** Cumulative incidence of TB among all participants enrolled after a Trace result (N=311)

| TB reference standard | Time (months) | TB (n) | Cumulative incidence of TB [%; 95% CI] | Competing events (n) | Remaining at risk (n) |
| --- | --- | --- | --- | --- | --- |
| CRS | 6 m | 150 | 49.0% [43.4 - 54.7%] | - | 112 |
|  | 12 m | 155 | 53.5% [46.9 - 60.1%] | - | 35 |
|  | 24 m | 156 | 54.9% [47.9 - 61.9%] | - | - |
| MRS | 6 m | 100 | 32.6% [27.4 - 37.8%] | 64 | 101 |
|  | 12 m | 106 | 37.3% [31.1 - 43.5%] | 64 | 31 |
|  | 24 m | 106 | 37.3% [31.1 - 43.5%] | 64 | - |
| Culture | 6 m | 68 | 22.0% [17.4 - 26.6%] | 79 | 112 |
|  | 12 m | 70 | 23.6% [18.5 - 28.7%] | 79 | 36 |
|  | 24 m | 70 | 23.6% [18.5 - 28.7%] | 81 | - |

**Figure 2.**
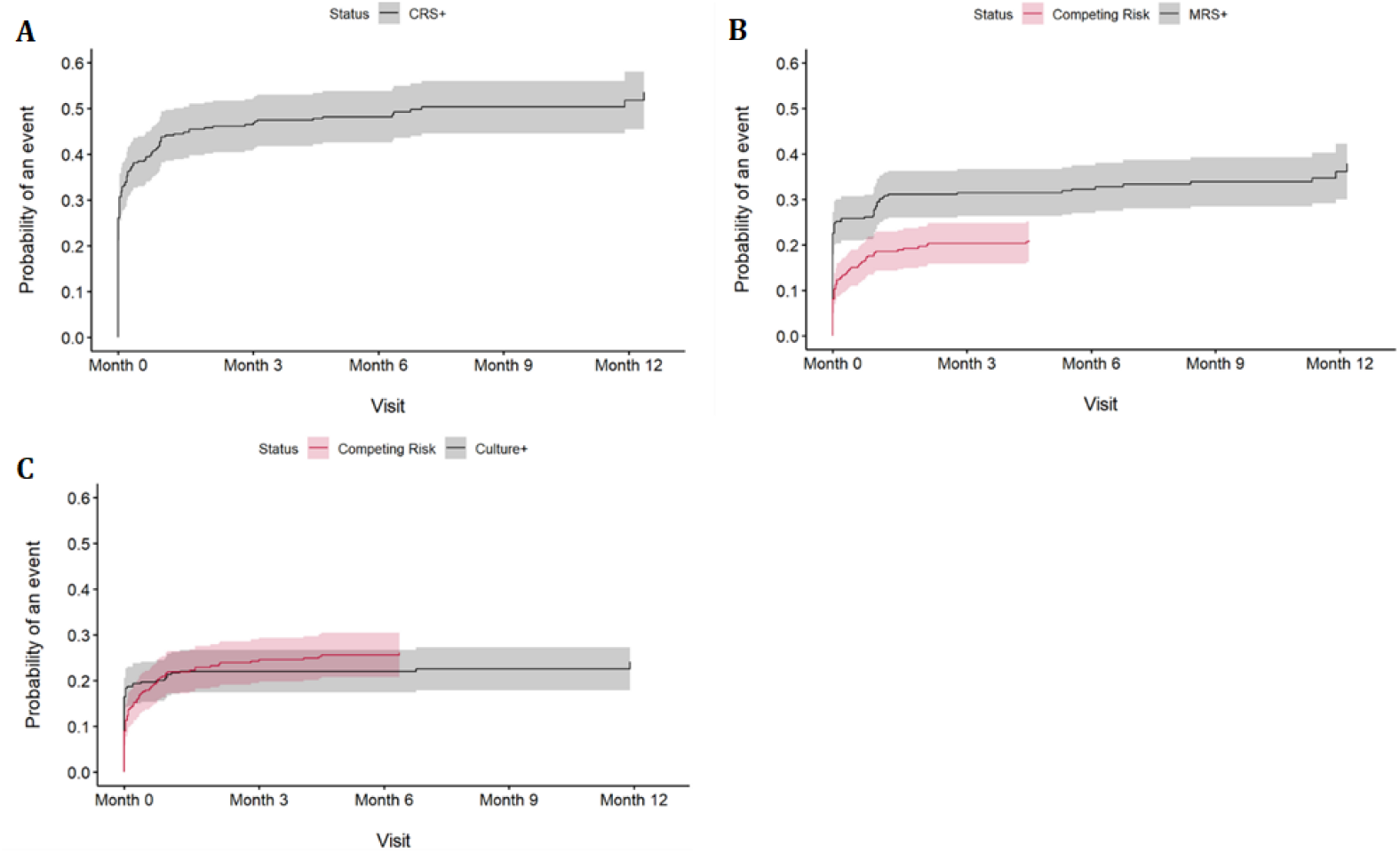
Cumulative incidence of TB, 0-12 months (full cohort): A) Composite reference standard (CRS); B) Microbiologic reference standard (MRS) C) Mtb culture positive. Cumulative incidence measured from baseline evaluation (black lines), with 95% confidence intervals (shading). Red line (and shading) corresponds to incidence of competing risk (initiation of TB treatment or death from TB) prior to event (culture-confirmed TB diagnosis).

### Cumulative incidence of diagnoses after baseline: Follow-up cohort

Among the 191 participants with negative TB status by CRS at baseline, cumulative incidence of TB defined by CRS was 16.7% (95% CI: 11.2 - 22.2%; n=30) by 6 months; 24% (95% CI: 15.6 - 32.4%; n=35) by 12 months; 26.4% (95% CI: 17% - 35.7%; n=36) by 24 months. Cumulative incidence of microbiologically confirmed TB was 16.7% (95% CI: 11.2 - 22.2%; n=30) at 6 months and 23.9% (95% CI: 15.9-31.9%; n=36) at 12 months (**Table 3**). For the same follow-up cohort over the same time periods, cumulative incidence of culture-positive TB was 3.8% (95% CI: 1-6.6%; n=7) at six months and 6.4% (95% CI: 1.6-11.2%; n=9) at 12 months. TB hazard rate was highest (CRS; **Figure 3**) for the first three-month interval following the baseline evaluation, and declined gradually over subsequent intervals (**Figure 3**).

**Table 3.** Cumulative incidence of TB in the Trace follow-up cohort (among participants who were TB-negative by CRS at baseline; N=191), followed through 24 months.

| TB reference standard | Time (months) | TB (n) | Cumulative incidence of TB [%; 95% CI] | Competing events (n) | Remaining at risk (n) |
| --- | --- | --- | --- | --- | --- |
| CRS | 6 m | 30 | 16.7% [11.2% - 22.2%] | - | - |
|  | 12 m | 35 | 24.0% [15.6% - 32.4%] | - | - |
|  | 24 m | 36 | 26.4% [17% - 35.7%] | - | - |
| MRS | 6 m | 30 | 16.4% [11% - 21.7%] | 15 | 101 |
|  | 12 m | 36 | 23.9% [15.9% - 31.9%] | 15 | 31 |
|  | 24 m | 36 | 23.9% [15.9% - 31.9%] | 15 | - |
| Culture | 6 m | 7 | 3.8% [1% - 6.6%] | 21 | 112 |
|  | 12 m | 9 | 6.4% [1.6% - 11.2%] | 21 | 36 |
|  | 24 m | 9 | 6.4% [1.6% - 11.2%] | 23 | - |

**Figure 3.**
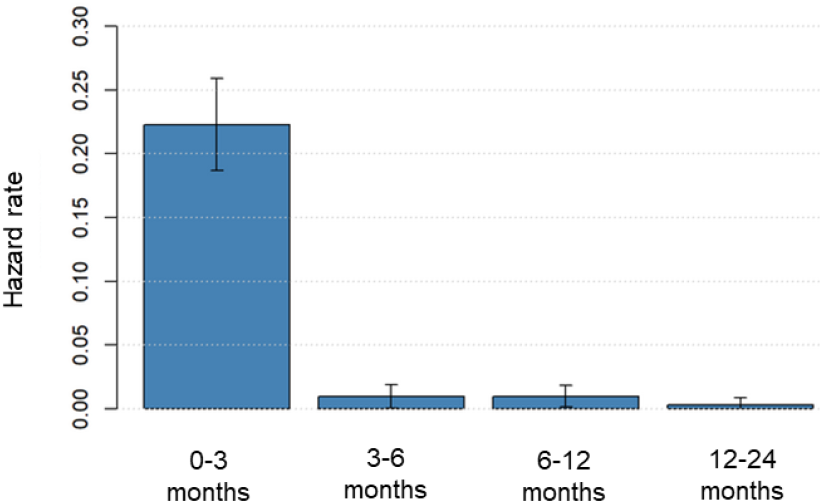
Estimated hazard rates for CRS-defined TB per month (excluding diagnoses made at baseline) across consecutive time intervals, with 95% confidence intervals. Hazard rates represent the instantaneous risk among those still event-free at the start of each interval. The highest hazard rate occurred in the first 3 months after baseline, followed by a gradual decline over subsequent intervals, reflecting continuing but decreasing event risk over time.

### Risk factors associated with hazard of TB

After exclusion of baseline diagnoses, advanced HIV status (with CD4 count <200) was a risk factor for microbiologically confirmed TB diagnosis within 1 year of a Trace result in the follow-up cohort (hazard ratio = 2.7 [95% CI: 0.9-8.0]), as was recent TB treatment (<5 years ago) compared to someone without recent TB treatment (hazard ratio = 2.6 [95% CI: 1.3-5.3]) (**Table 4**). Considering ancillary baseline diagnostic testing, both abnormal chest x-ray (found in 65% of the cohort) and positive tongue swab (found in only 9%) were also associated with higher risk of TB in the follow-up cohort (hazard ratios 2.4 [95% CI: 1.1-5.2] and 4.5 [95% CI: 1.3-15.2], respectively).

**Table 4.** Relative hazard of microbiologically confirmed TB (MRS; 0-12 months follow-up) comparing baseline characteristics and test results, both including (total cohort) and excluding (follow-up cohort) diagnoses (CRS) made at baseline evaluation. Results from univariable Cox proportional hazards models; hazard ratios (HRs) represent the relative risk of the event associated with the comparator group (gray) relative to the reference group (white).

|  | Baseline characteristics | Total cohort (N=311) |  |  |  | Follow-up cohort (N=191) |  |  |  |
| --- | --- | --- | --- | --- | --- | --- | --- | --- | --- |
|  |  | *n* * | (%) | HR | (95% CI) | *n* * | (%) | HR | (95% CI) |
| **Age** | < 35 | 128 | (41%) | 0.7 | (0.5-1) | 71 | (37%) | 1.3 | (0.6-2.6) |
| **HIV Status** | Negative | 133/301 | (44%) | (ref) |  | 92/189 | (49%) |  |  |
|  | HIV+ CD4 ≤200 | 35/301 | (12%) | **2.8** | **(1.6-4.7)** | 12/189 | (6%) | 2.7 | (0.9-8.0) |
|  | HIV+ CD4 > 200 | 133/301 | (44%) | 1.0 | (0.7-1.6) | 85/189 | (45%) | 1.1 | (0.6-2.3) |
| **Recent TB treatment** | No | 197/305 | (65%) | (ref) |  | 107/189 | (57%) |  |  |
|  | <5 years | 72/305 | (24%) | 0.8 | (0.5-1.2) | 58/189 | (31%) | **2.6** | **(1.3-5.3)** |
|  | >5 years | 36/305 | (12%) | 0.5 | (0.2-1.0) | 24/189 | (13%) | 0.9 | (0.3-3.1) |
| **Country** | S. Africa | 141 | (45%) | (ref) |  | 100 | (52%) |  |  |
|  | Uganda | 170 | (55%) | 1.0 | (0.7-1.5) | **91** | **(48%)** | **0.3** | **(0.2-0.7)** |
| **Sex** | Male | 155 | (50%) | (ref) |  | 83 | (43%) |  |  |
|  | Female | 156 | (50%) | 0.7 | (0.5-1.0) | **108** | **(57%)** | **0.5** | **(0.2-0.9)** |
| **Baseline chest X-Ray result** | Normal | 106/302 | (35%) | (ref) |  | 77/184 | (42%) |  |  |
|  | Abnormal | 196/302 | (65%) | **1.7** | **(1.1-2.6)** | **107/184** | **(58%)** | **2.4** | **(1.1-5.2)** |
| **Baseline tongue swab result** | Negative | 201/220 | (91%) | (ref) |  | 132/138 | (96%) |  |  |
|  | Positive | 19/220 | (9%) | **4.5** | **(2.5-8.0)** | **6/138** | **(4%)** | **4.5** | **(1.3-15.2)** |

\*Denominators are presented where complete data were not available, otherwise N=311/ N=191, as indicated at top of table

## Discussion

Among individuals with a sputum Xpert Trace result during routine care, cumulative incidence of TB within 12 months in the overall cohort was 24% by Mtb culture, 37% by MRS, and 54% by CRS. After 12 months, TB incidence did not increase for culture and MRS, but had a 1% increase using the CRS definition, though the sample size in this later time period was small. While most diagnoses resulted from the baseline evaluation performed shortly after the Trace result, we also observed 6% cumulative incidence for culture-confirmed TB and 24% by MRS and CRS definitions within 12 months, among those without a baseline TB diagnosis. These estimates quantify how much TB disease would be missed if Trace results were interpreted as ‘no TB disease,’ either as a stand-alone test (overall cohort) or after a full clinical evaluation including results of Xpert re-testing and culture (follow-up cohort).

While TB has previously been classified in two distinct stages (latent and active), it is better characterized as a continuum of infection and disease states.^19^ Moreover, among individuals who develop TB disease, progression can be rapid or gradual, and may fluctuate between periods of progression and partial recovery. Trace results may arise when TB hovers near or recedes below the limit of culture detection, and such patients may still have active disease and remain at risk for further disease progression. In our cohort, diagnoses continued to occur more than three months after participants’ initial sputum Trace result, even after multiple negative sputum tests and negative TB evaluations by expert clinicians. However, roughly half of participants were monitored off of TB therapy without developing further microbiological or clinical evidence of TB disease.

Our estimates of the TB burden among people with Trace diagnostic results are similar to estimates from a recent study of individuals who received a Trace result during a symptom-agnostic community screening program^15^. A similar proportion were diagnosed at baseline (35% in the community screening cohort; 39% in this clinical cohort), and cumulative incidence of TB diagnoses by 24 months, excluding baseline diagnoses, was nearly equal (28% vs. 26%, respectively– **Table S-2**).

At least two baseline tests were found to be associated with an increased hazard of TB disease diagnosis within a year following baseline clinical evaluation. Abnormal chest x-ray results had a modest prognostic value, with a 2.4 times greater risk of microbiologically confirmed TB. A similar result was observed in the community cohort, but the magnitude was larger, perhaps due to a lower proportion of people living with or prior TB. The results of tongue swabs were also modest. Although the small proportion of participants (9%) with positive tongue swab results had a 4.5-fold increased risk of microbiologically confirmed TB compared to those who tested negative, the swab results had a low positive predictive value. Tongue swabs could instead be useful for identifying a subset of individuals at considerably higher risk of progressive disease, with high specificity. Since tongue swabs are most sensitive among people with higher semiquantitative sputum Xpert results^18^, they may be detecting the subset of people with Trace results who have more Mtb DNA in their respiratory tract.

Among participants with a Trace positive result who were retested by Xpert Ultra at baseline, 28% of participants had a semiquantitative result greater than Trace upon retest in this cohort, which illustrates the fluctuation of Mtb DNA quantified in sputum specimens from one sample point to the next. Non-sputum diagnostics like chest x-ray and tongue swabs may be useful for risk stratifying which patients with a Trace result are most likely to benefit from TB treatment, but a more comprehensive risk algorithm will be required.

This study had several strengths and limitations. We used a prospective cohort design with comprehensive clinical and diagnostic follow-up of Trace positive individuals. Our estimates of cumulative disease burden account for both censoring and competing risks using standard methods for participants who initiated TB treatment during follow-up. However, these methods assume competing events are non-informative, which may not hold since participants recommended for therapy could be at higher risk for developing microbiologically confirmed TB disease (i.e., related to the clinical judgement recommend therapy). If the non-informative censoring assumption were incorrect, then our results would be underestimating TB disease in this population. Nevertheless, accounting for competing risks using standard methods provides more accurate estimates than ignoring these events entirely, which would lead to even greater bias.

Our study design lacked a parallel comparison group of individuals with a negative sputum Ultra, which would have allowed a determination of background TB incidence rates experienced by our cohort. Without sequencing to identify reinfection, we cannot definitively distinguish between progression of early tuberculosis, delayed detection of prevalent disease below baseline detection thresholds, and new infections acquired during the study period. While this is the largest longitudinal cohort of participants with a Trace result to date, some of our analytic subgroups stratified by baseline characteristics were relatively small, resulting in cumulative incidence and hazard ratios with wide confidence intervals. Relatedly, we also limited our analysis to univariate analysis of risk factors, which may obscure stronger associations of certain factors in combination (e.g., CXR and symptoms may have been stronger predictors among people without prior TB).

In conclusion, our results suggest that among people with a Trace sputum result from routine clinical care, roughly one-quarter have microbiological confirmation of Mtb and over half will be started on treatment within a year. The paucibacillary disease represented by Trace results includes disease that is otherwise microbiologically undetectable for more than three months despite repeated clinical and sputum diagnostic evaluations. Beyond their clinical implications, these findings support the inclusion of Trace test results in the assessment of new diagnostic tools rather than excluding them as though indeterminate. Even among Trace positive individuals who are not diagnosed with TB after extensive workup and follow-up, our estimates suggest an ongoing, though diminishing, risk of disease for at least one year.

**Supplementary Table S1:**
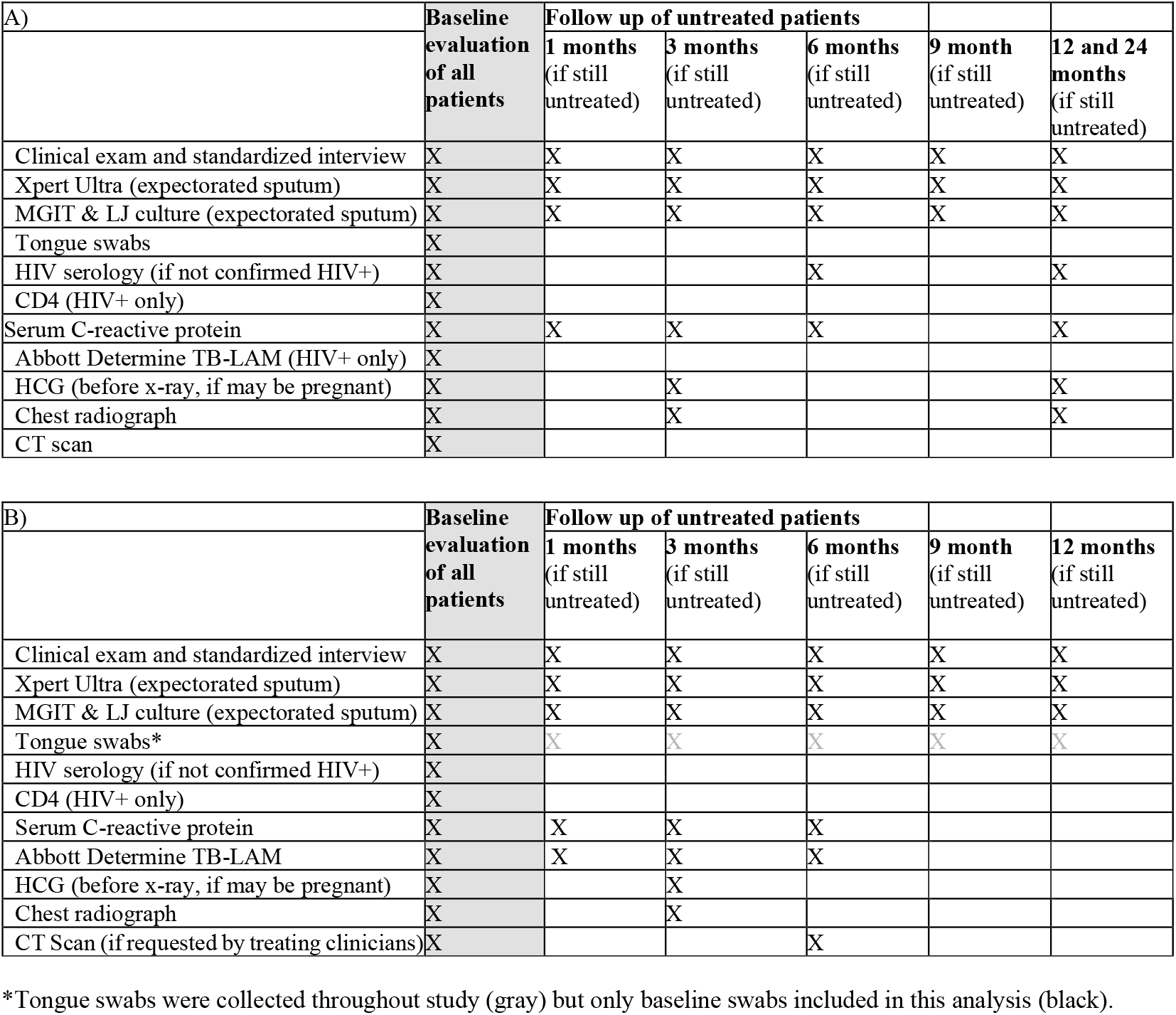
Table of procedures per site: A) schedule used in Uganda B) schedule used in South Africa.

**Supplementary Table S2:**
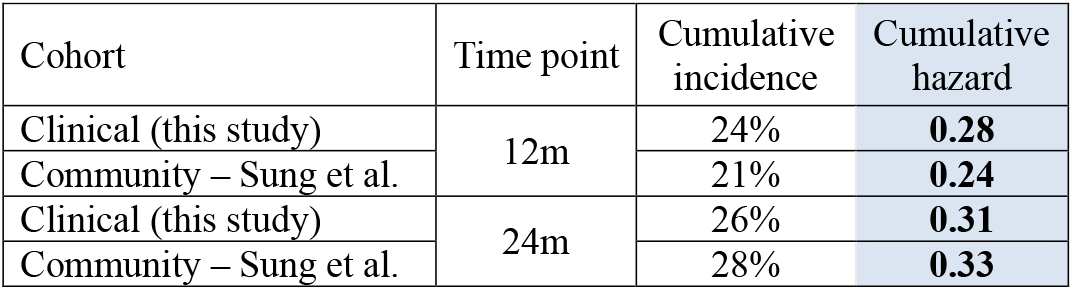
Comparison of cumulative hazard of a TB diagnosis after Trace result follow-up cohorts where baseline diagnoses are excluded from a community cohort (Sung et al., 2025) vs. this clinical cohort. In the community cohort, TB diagnosis was defined as a recommendation for treatment. In this clinical cohort, TB diagnosis was defined as a recommendation for treatment or a culture-positive result or TB-related death, however the resulting data showed the categories to be functionally equivalent. Our estimates of cumulative incidence were converted to cumulative hazards, in order to directly compare to those observed in a recently published community cohort^*15*^, via the relationship between cumulative incidence and cumulative hazard: *Cumulative hazard =−log(1−cumulative incidence)*

## Data availability

The deidentified dataset used for this study and a data dictionary are being uploaded to a controlled access data archive, per requirements of the institutional review board that reviewed and approved this study. The upload process is under review and this preprint will be updated with a link to the data as soon as available. At this time, all data produced in the present study are available upon reasonable request to the authors

## Acknowledgements

This work would not be possible without the support of the staff of Harry Gwala Regional Hospital and referring primary health care clinics, the KwaZulu-Natal Department of Health, the South African National Health Laboratory Service, the TURN-TB research team and physician consultants, and the clinical and laboratory staff at Kitebi Health Centre, Kisugu Health Centre, Kisenyi Health Centre, Kawaala Health Centre, Kiswa Health Centre, Naguru Hospital, and Mulago Hospital Wards 5&6.

